# Computational fluid dynamic (CFD), air flow-droplet dispersion, and indoor CO_2_ analysis for healthy public space configuration to comply with COVID 19 protocol

**DOI:** 10.1101/2020.07.02.20145219

**Authors:** Andrio Adwibowo

**Affiliations:** U. o. Indonesia., West Java, Indonesia

**Keywords:** air flow, droplet, distance, CFD, LBM

## Abstract

The droplet has a limited travel distance. Nonetheless, especially in the indoor public space the air flow can propagate the droplet to travel long distance. Based on this situation, this paper aims to study the relationships of seat configuration-social distance-air flow-droplet dispersions. The analysis was based on the computational fluid dynamic (CFD) using lattice-Boltzmann model (LBM). The result confirms that by modifying public space configuration in this case by providing more space and increasing seating distance can reduce the vulnerability towards droplet dispersions. Whereas, providing shield including adding protection is far more effective in avoiding dispersions. The public space reconfiguration including increasing seat distance and reducing seating capacity also has an effect in reducing the indoor CO_2_. Capacity reduction from full capacity to 30% can decrease the CO_2_ from 5722 to 2144 ppm.

## Introduction

Droplets and aerosols formed in the respiratory tract or also known as via virus-laden fluid particles are facilitating the respiratory infection transmission. This droplet is expelled from the mouth and nose during breathing, talking, coughing, and sneezing. Large droplets expelled with sufficient momentum can directly impact the recipients’ mouth, nose, and conjunctiva.

The infection transmission process is mediated by complex flow phenomena including physical factors namely turbulent jets and flow-induced particle dispersion. Since the transmission is function of physical properties, then flow physics is central to study the transmission.

The public spaces including classroom, restaurant, and even transportation cabin are experiencing flow phenomena and this make those places prone to the droplet dispersions. A research by Lu *et al*. (2020) have studied how the crowded public space in this case a restaurant can be a potential space of droplet dispersion facilitated by air conditioned ventilation system.

Virus transmission is a function of environment attributes including air flow. Meaning to say, the transmission cannot be explained by droplet dispersion alone. Whereas, the droplet has dispersion limitations and travel in short distance only. Larger respiratory droplets (>5 μm) remain in the air for only a short time and travel only short distances generally <1 m. In fact according to droplet dispersion in a restaurant, the distance of infected person and persons at other tables were all >1 m beyond the travel distance range of the droplets. Whereas, the droplet can travel long distance >1 m because it was propagated by the air flow from the air conditioned ventilation system.

Following the restaurant based dispersion case discussed above, Li *et al*. (2020) have applied CFD to model the correspondent air flow and dispersion. Based on the model, they found that the dispersion was related to air flow, crowd, and even seating duration in a space when the infected person was present.

Most of current studies on the relationships of people-seating configuration-air flow-droplet dispersion were not based on model yet. If there is a model, a modified model based scenario including increasing the seating distance or even adding protective structure is still limited. Likewise, this paper aims to model the relationships of people-seating configuration-air flow-droplet dispersion in indoor public space settings. Air flow model used in this study is based on computational fluid dynamic (CFD) analysis.

## Methodology

The analysis in this study consist 2 steps. First analysis emphasized on air flow that contain droplets is modeled using Computational Fluid Dynamic (CFD). This aims to see the flow behavior across the various seating configuration settings in a room. The second analysis is calculating the air quality based on CO_2_ model for every seating configuration and capacity.

### Computational Fluid Dynamic

CFD using in this study is a two dimensional flow model. Initially the particle is flowing across a linear barrier and in this study the barriers are the seating configuration. The presence of chair and added with the protective shield is hypothetically can divert the flow and create vortices. A control and intervention scenarios have been prepared to analyze whether the seating scenarios can reduce the spread of droplets.

The control is the default seating configuration without shield and no distance as well. While, the scenario includes the seating configuration with distance and seating arrangement provided with protective shield.

The CFD model of particle flows across the barriers is using the lattice-Boltzmann method (LBM). This method has been widely used to study the air flow and suitable for indoor setting (Crouse *et al*. 2002, Khan *et al*. 2015). The LBM based on threefold discretization of the Boltzmann equation including phase space, involving space, time, and velocities. The movement and distributions of a flow particle are described by particle distribution functions residing at the sites of a regular grid (lattice) of points which encompasses an entire indoor environment including a room. The particle distribution functions represent the particle presence probability with a given velocity at each lattice site. The lattice is given as follow equation:

Using model in Figure 1, a discretize two-dimensional space with a square lattice is developed. In this model, 9 fundamental displacements and velocities were also developed. The model was completed using simulation variables *n*_*i*_ consist of 9 densities at each lattice of particles with the 9 allowed velocities. Then, a total density ρ and macroscopic flow velocity *µ* are denoted as:

**Figure 1.**
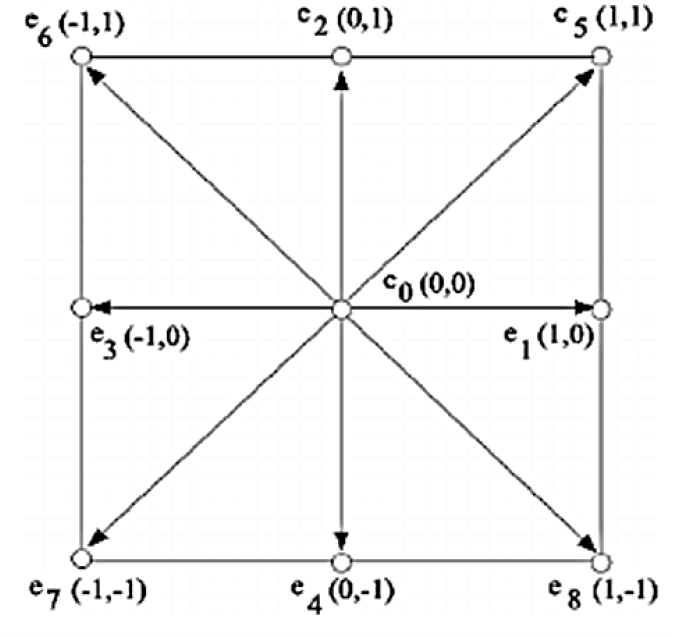
The Lattice-Boltzmann model.

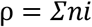

Particle in axis x is denoted as:

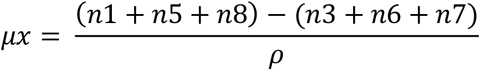

Particle in axis y is denoted as:

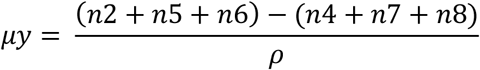

## Results

Figure 2,3, and 4 present the air flow across the seating configuration. The air flow direction was modeled from the front side. There were 3 scenarios were developed, they were seating configuration without distance and protective structure, with distance, and with protective structure. The seating configuration was classroom configuration with several rows. This is the general configuration that is commonly used in the school, teaching theatre, and even transportation facility including bus and plane.

**Figure 2.**
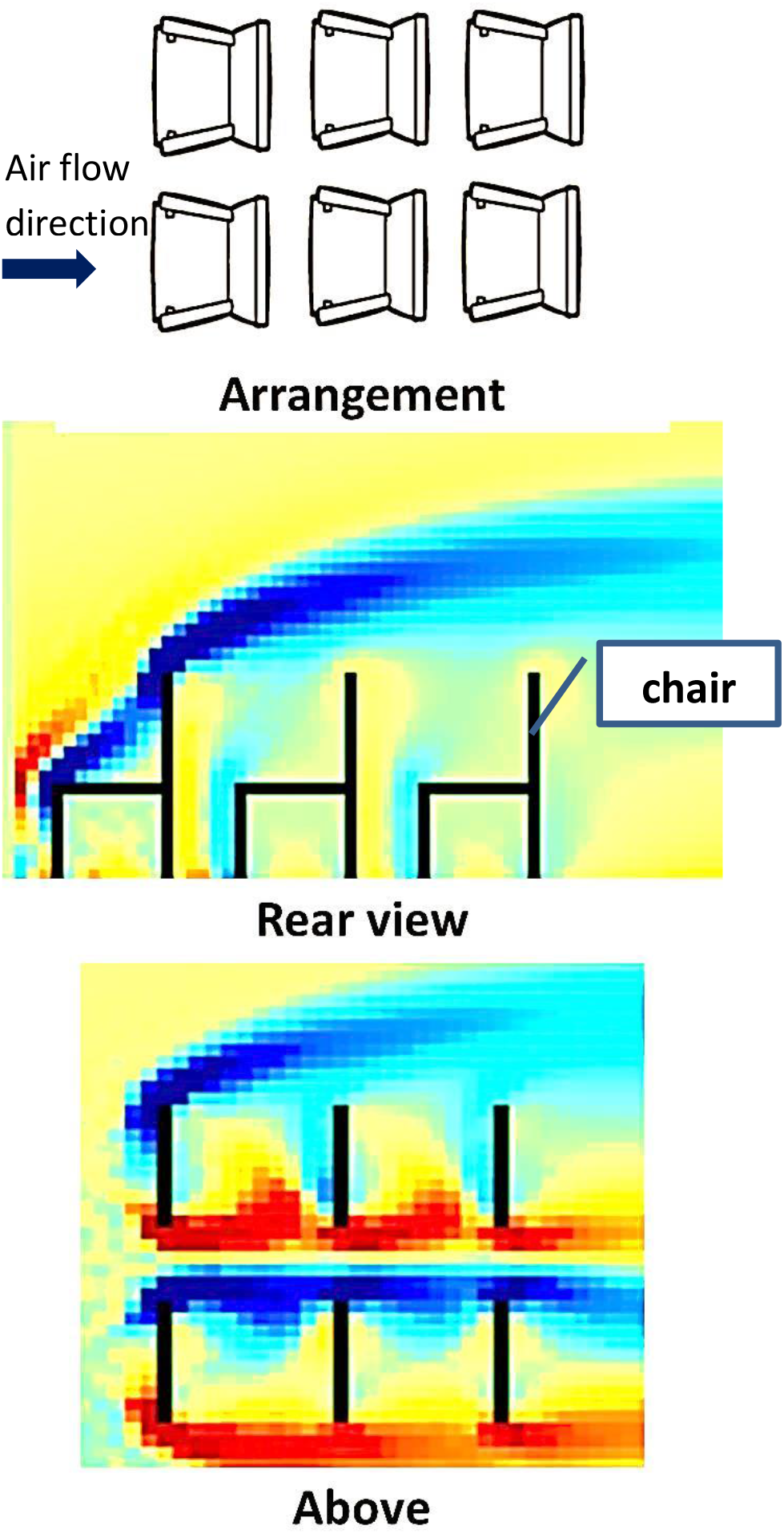
CFD of air flow and droplet dispersion (blue area) without seating arrangement/seat configuration (without distance).

**Figure 3.**
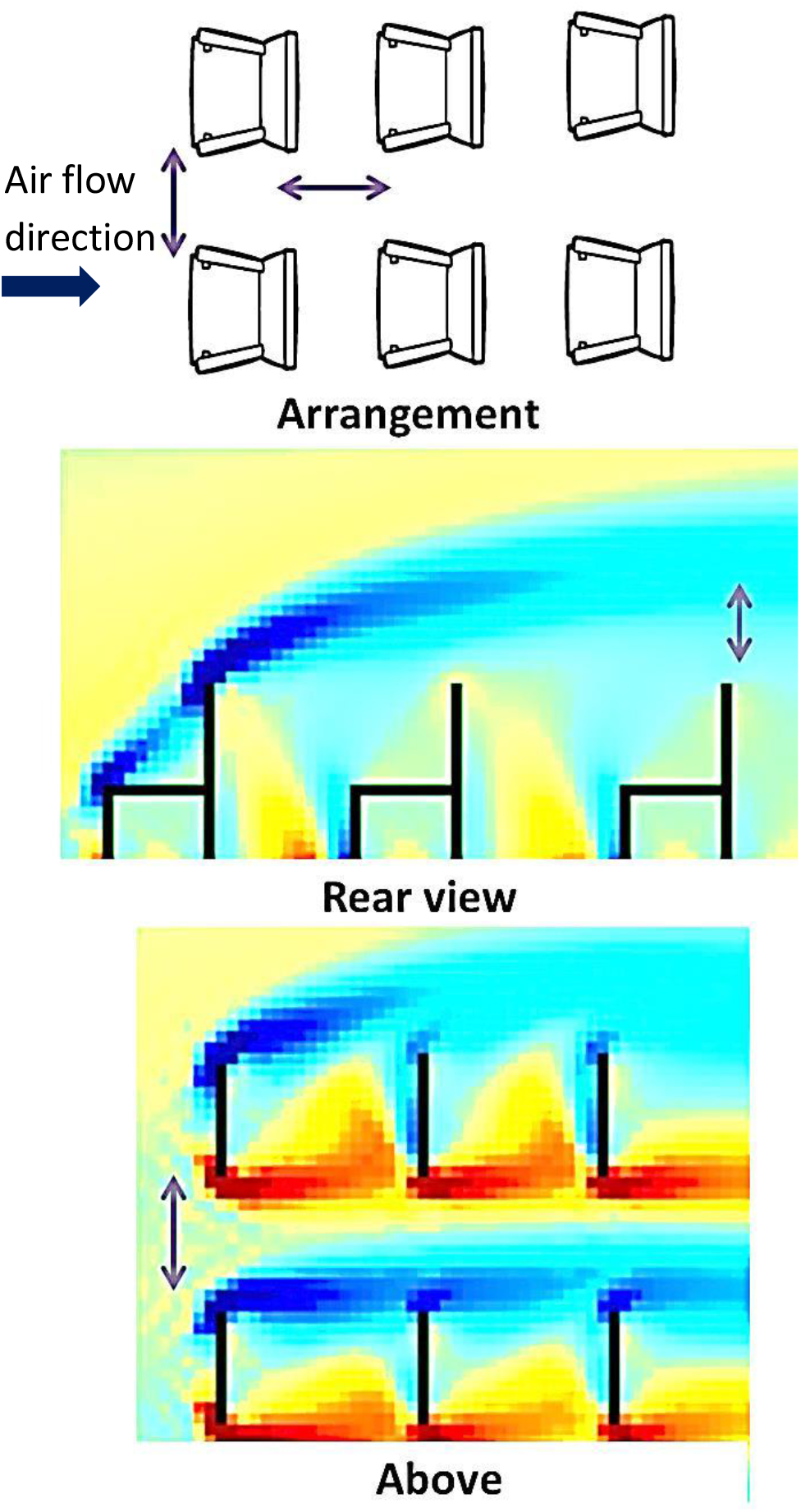
CFD of air flow and droplet dispersion (blue area) with seating arrangement/seat configuration (with distance).

**Figure 4.**
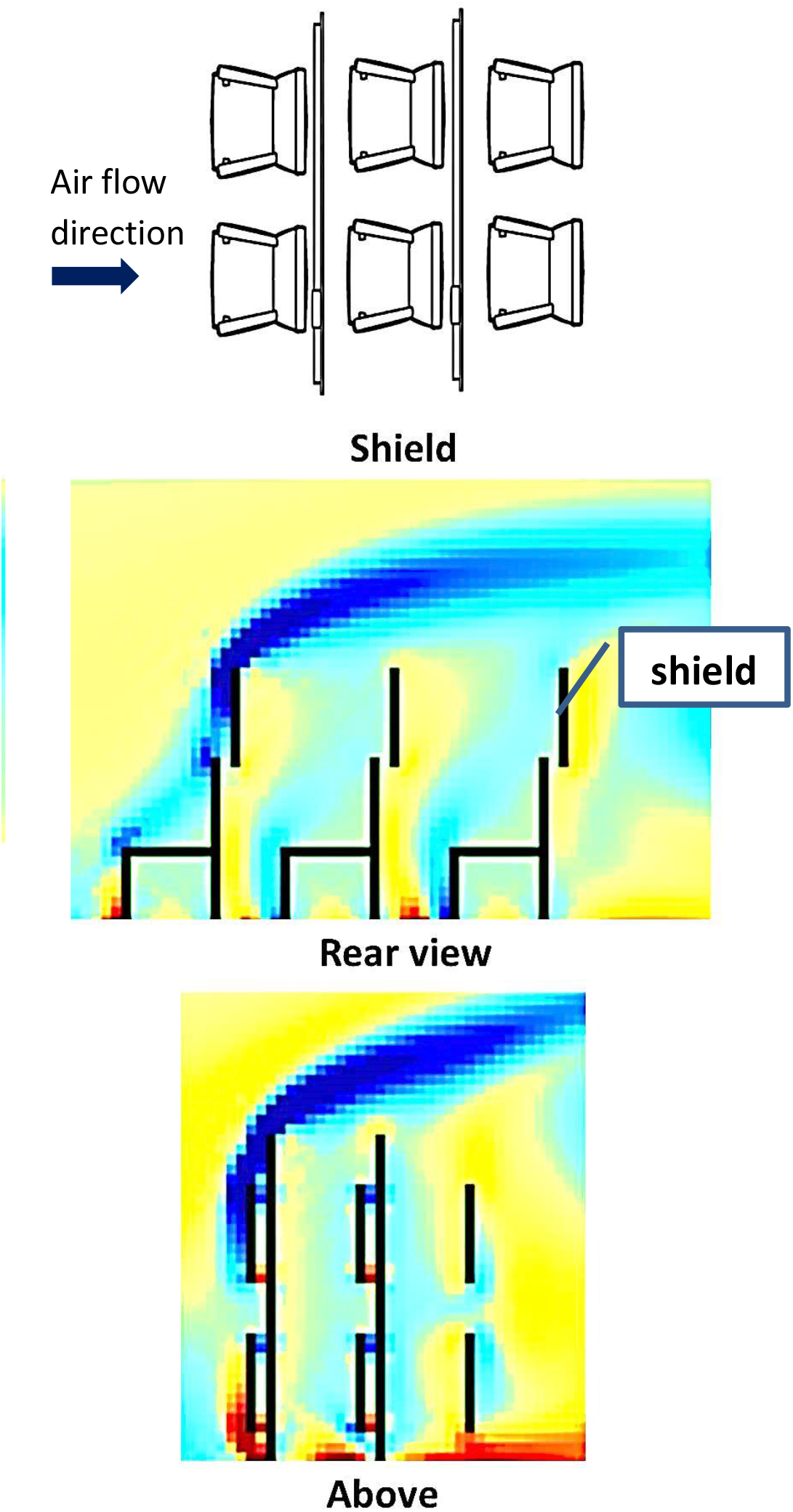
CFD of air flow and droplet dispersion (blue area) with overhead shield protection installed on chair.

In the first scenario (Figure 2), it shows that the air flow can reach and very close to the second and third rows. The second and third rows were arranged at standard distance.

The second scenario (Figure 3) shows that there was an increased distance between rows. As a result, there was a space between the air flow and second row and it was obvious for the third row.

The third scenario (Figure 4) was the scenario when there is no distance between rows. Nonetheless, a shield was installed to border row. The aim of shield installment is to observe whether the shield can obstruct the air flow. The results showed that the second and third rows were free from air flow. The shield can obstruct the air flow and the air is flowing above the second and third rows.

This paper also tested the indoor CO_2_ production under seat capacity. Increasing seating distance means reducing number of seat available in a classroom. Figure 5 and 6 shows the indoor CO_2_ production under different numbers of seating capacity (30% to full) and outdoor CO_2_. It clears that the indoor CO_2_ has positive correlation with the seat capacity. Reducing seating capacity can reduce the indoor CO_2_.

**Figure 5.**
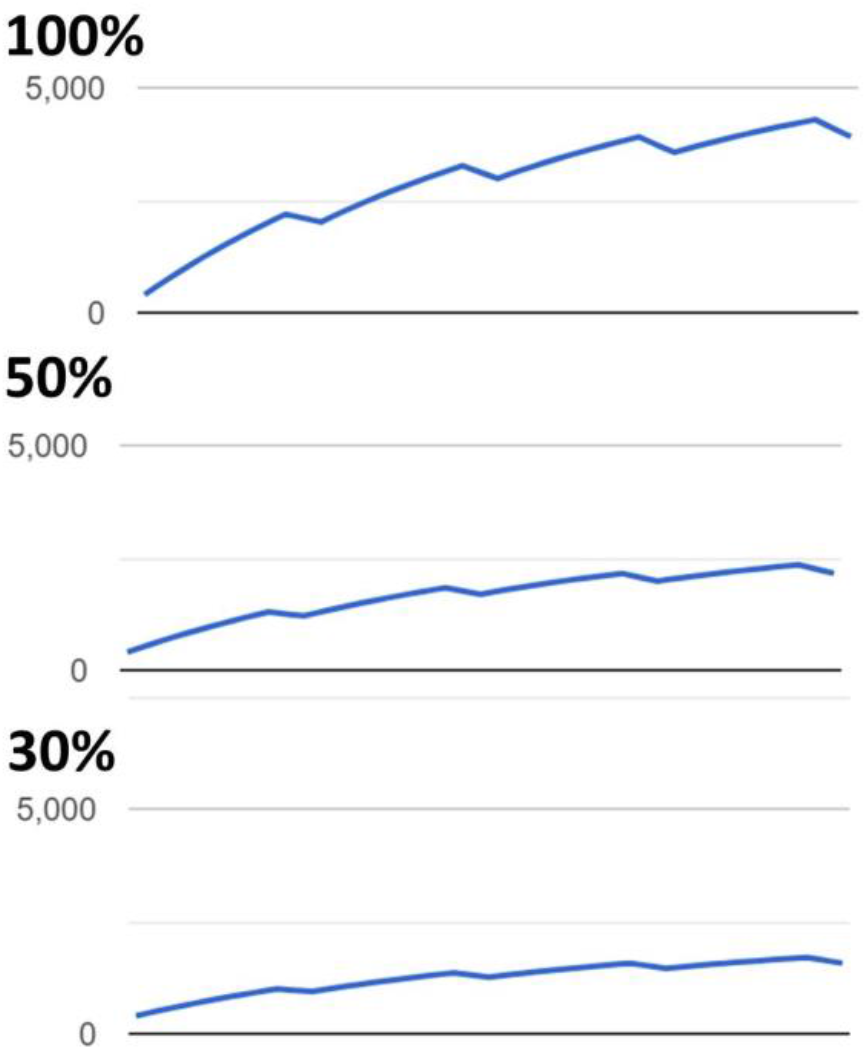
Indoor CO_2_ (ppm) with full, 50%, and 30% seating capacity.

**Figure 6.**
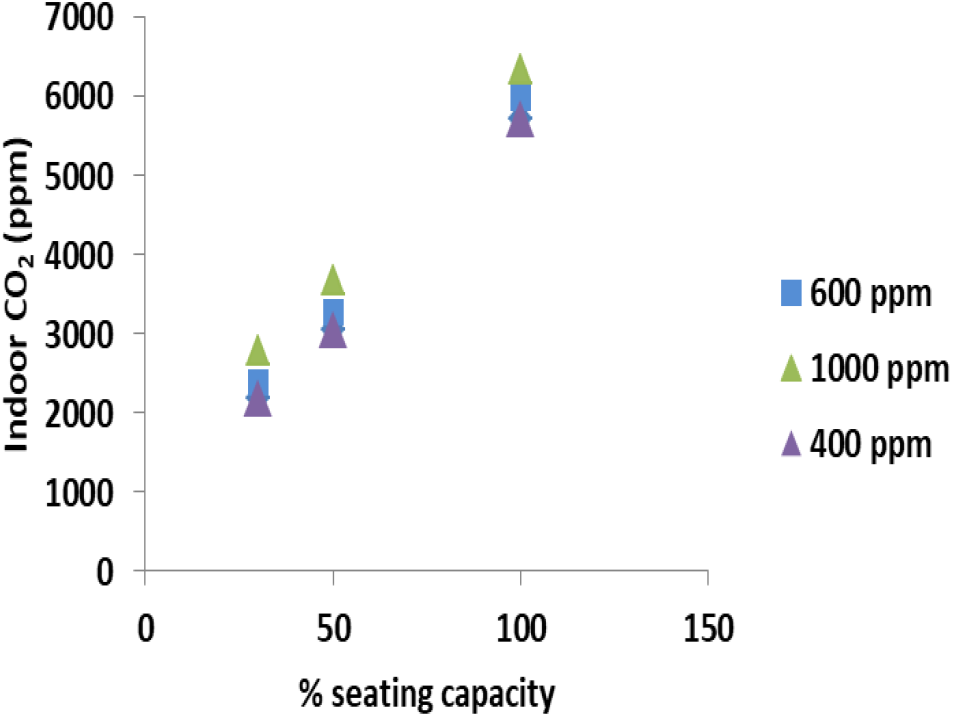
Effects of seating capacity on Indoor CO_2_ (ppm) under different outdoor CO_2_ (400 – 1000 ppm)

## Discussion

Public spaces arrangement and safety have been concerned regarding the potential COVID 19 spread inside the public spaces, including classroom, restaurant, and even transportation cabin. Some measures have been recommended including social distancing practice. In this context, it is advised to spread out desks and chairs to at least a couple of meters (0.9-1.5 m) between each station and followed by the occupancy of number of guests per time to a minimum. Another issue is to maintain the heating, ventilation, and air condition system and make sure any filtration system to work properly combines with regular cleaning and maintenance.

The distance effects on public spaces whether in class room, restaurant, or transportation play fundamental roles in droplet dispersions. One of public space that vulnerable is the educational institution ranging from kindergarten, elementary school, high school to campus. This situation related to the number of occupancy rates per hour. A classroom can be occupied with 100 students and they are sitting very close each other and experiencing overcrowding as well. To anticipate this, seating arrangement should provide a space to ensure from 0.9 to 1.5 m distance from other students as required by COVID 19 protocol (Abale, Charak 2020).

A behavior and dynamic of air flow with its particle contained have led to numerous researches using CFD approaches. A CFD work to investigate the indoor air quality has been pioneered by Yang *et al* (2003). In their paper they have modeled how the air conditioning system hanged on the wall can undertake indoor heat load and conduct good indoor thermal comfort. The CFD has succeeded to model the air flow, wind velocity, and behavior in activity area where people sit and where the people can feel the wind flow. The most advantage is the model has indicated local areas without ventilation where toxic gases not discharged in time.

The notorious of CFD is its ability to model the air flow behavior (Huang and Lin 2014, Pamonpol *et al*. 2020, Zhang *et al*. 2016). The air flow becomes important issues since the air may carry and transport hazardous particles including virus particles. According to previous study taken example in restaurant, a transmission was related to the seating arrangement and air flow direction released from nearby air conditioned ventilation unit. A restaurant cluster was observed in 9 people who had been sitting at other tables several metres away near air ventilation. In an indoor space, all people shared one thing in common which is an air from singular source.

The transmission risk and droplet dispersion models of seating configuration in public space without distance and even without protection have been clarified through model by team from U. of Oregon. In one model, a droplet released by infected persons and caught in a current of air coming from an air conditioning unit was carried by the air to people at other tables. This model indicates that the air flow released from ventilation unit can be distributed to other areas. In indoor settings, virus transmission could not have been by respiratory droplet transmission alone, whereas a stronger air flow produced from air conditioned ventilation unit is also responsible to spread the droplets (Ezratty and Squinazi 2008, Prussin *et al*. 2016, Goyal *et al*. 2008).

A crowded place with no distance seating configuration is also an evidence of dispersion in indoor public space. According to Park *et al*. (2020), droplet dispersion and infection in a call center occupied by 216 persons was due to a high-density work environment and can become a high-risk site. In fact 94 persons out of 216 persons that were seating closed each other were infected.

This study clarifies the advantage of protection that is hypothetically more efficient to provide protection from COVID 19. In this study based on the CFD model, providing shield when seating is proven more effective in providing protection. Mittal *et al*. (2020) have discussed comprehensively covering the protection based mitigation measures. Basically the shield is categorized as inward protection. In comparison to mask, mask can be categorized as both inward and outward protection since it is wear in the mouth. As inward protection, the shield can provide structure that can block the aerosolized particles that would otherwise be inhaled.

Recently, shield like structure has been considered as versatile solution when the social distance and seating configuration are difficult to implement. Seating arrangement in the plane can be an example how increasing seating distance cannot be the option. Airplane manufacturer already equipped the seat with this shield like structure. Even in classroom in some countries already installed a shield screen to protect the students.

This study confirms an effective blockage of air flow following shield installment and this comparable to result from another study. The effectiveness of a protective structure to influence the air flow has been studied by Hasan *et al*. (2016). In their study, a structure in the form of partition placed between beds in health care ward can reduce and even block the air flow (Figure 7).

**Figure 7.**
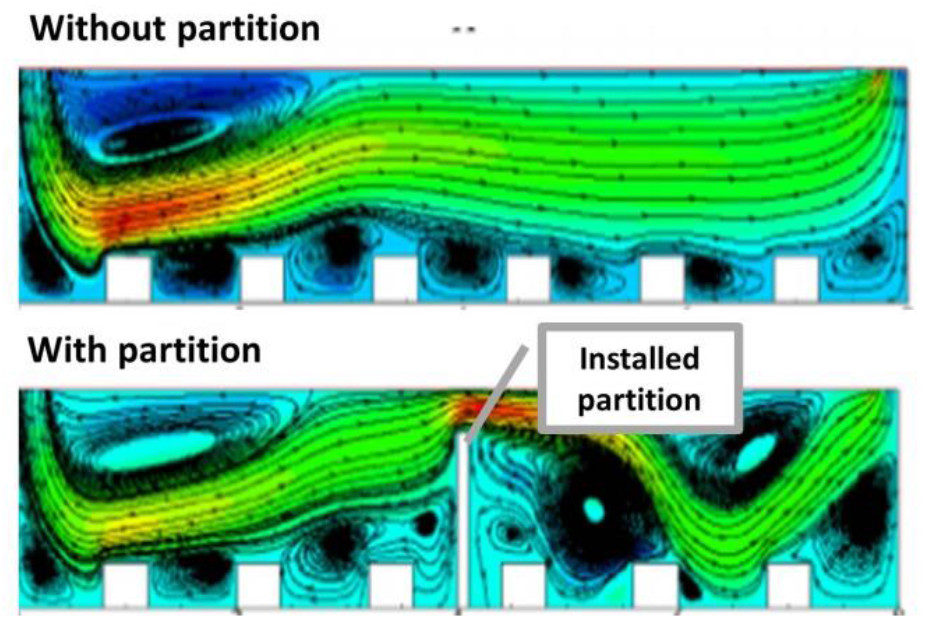
CFD of air flow with partition installed between beds (Hasan *et al*. 2016).

## Conclusion

It has been discussed that the social distancing and wearing protection including face mask and shield can reduce the vulnerability to droplet dispersion. Likewise, this paper has provided an empirical study how those measures can be effective in avoiding the droplet dispersion. In a classroom setting, maintaining seating configuration by reducing seating capacity and even adding protective structure can be an effective measure to reduce and avoid droplet dispersions and even increase air quality.

## Recommendations

Modifying the current public space configuration and capacity can be an option to effectively reduce the droplet dispersion and transmission rates. Following to that, it is recommended that the decision and strategy regarding the public space arrangement should be based on empirical study using robust model and CFD can be the option.

## Data Availability

The data are available in manuscript

